# Antibody Response to CoronaVac Vaccine in Indonesian COVID-19 Survivor

**DOI:** 10.1101/2021.05.28.21254613

**Authors:** Rahmat Azhari Kemal, Dita Kartika Sari, Ariza Julia Paulina

## Abstract

Several studies have shown that individuals with previous history of SARS-CoV-2 infection had boosted antibody response after single dose of mRNA or adenovirus-vectored vaccines. We wondered whether single dose CoronaVac, a whole-inactivated vaccine, could be considered for COVID-19 survivors in Indonesia. We measured IgG anti-RBD titre among 18 survivors and 37 non-survivors. Among survivors, there were 9 survivors with positive antibody titre (seropositive) before vaccination and 9 seronegative survivors. All respondents received two doses of CoronaVac vaccine at 14-days interval. We found no significant antibody titre difference between non-survivor at 14 or 28 days after second dose as well as seronegative survivor at at 14 days after second dose. Seropositive survivors were rapidly boosted after first dose with higher antibody titer than non-survivors and seronegative survivors after second dose. However, antibody titer did not differ between first and second dose among seropositive survivors. Seropositive COVID-19 survivors could receive single dose of CoronaVac vaccine which could potentially ease the vaccine supply constrain. A long-term follow-up must be conducted to observe difference in antibody response and persistence.

**Article Summary Line:** Seropositive COVID-19 survivors had significantly higher antibody response after first dose of CoronaVac vaccine compared to non-survivors and seronegative survivors after second dose of vaccine.

## MANUSCRIPT

Several studies have assessed antibody response to mRNA and adenovirus-vectored vaccines in COVID-19 survivor. All studies shown that individuals with previous history of SARS-CoV-2 infection mounted significantly higher post-vaccination antibody compared to naïve individuals.^1-4^ Indonesia has launched its COVID-19 vaccination programme since January 13^th^ 2021 utilising whole-inactivated vaccine, CoronaVac (Sinovac, China). Initially, COVID-19 survivor was not recommended for vaccinated. However, since February 2021, COVID-19 survivor could be vaccinated at least 3 months after PCR confirmation. Due to constrain in global COVID-19 vaccine supply, we wondered whether single dose whole-inactivated vaccine could be considered for COVID-19 survivors in Indonesia.

We conducted preliminary studies among participants with or without prior PCR-confirmed SARS-CoV-2 infection. There were 18 survivors and 37 non-survivors. All participants had CoronaVac vaccination with 14-days interval as initially assigned for adults 18-59 years old. For IgG anti-RBD measurement, 23 non-survivors were sampled at 14 days after second dose (day 28), 13 non-survivors were sampled at 28 days after second dose (day 42) and one non-survivor was sampled at days 28 and 42. Blood samples from six survivors were taken at days 0, 14, 28 of vaccination. One survivor was also sampled at day 42. Three survivors were only sampled at days 0 and 14. IgG anti-RBD was measured from serum using Elecsys® (Roche Diagnostics). The study was approved by Ethical Committee of the Faculty of Medicine Universitas Riau (No: B/023/UN19.5.1.1.8/UEPPK/2021)

Demographical data of respondents is presented in Table 1. There was a significant difference on COVID duration between seronegative and seropositive survivors, with seropositive survivors had longer duration. There was no significant difference on distance between post-COVID and vaccination time between both groups.

**Table 1.** Demographical data of respondents

| **Groups** | **n** | **Female**<br>**n (%)** | **Age, years**<br>**Median (Min-**<br>**Max)** | **COVID-19**<br>**length, days**<br>**Mean ± SD** | **Time post**<br>**COVID-19, days**<br>**Mean ± SD** |
| --- | --- | --- | --- | --- | --- |
| **Total** | 55 | 35 (63.7%) | 25 (21 – 51) | N/A | N/A |
| **Non-survivor\*** |  |  |  |  |  |
| Day 28 | 24 | 18 (75%) | 24.5 (21 – 48) | N/A | N/A |
| Day 42 | 14 | 9 (64.3%) | 29.5 (22 – 51) | N/A | N/A |
| **Survivor** |  |  |  |  |  |
| Seronegative | 9 | 5 (55.6%) | 23 (22 – 43) | 3.3 ± 1.7 | 166.4 ± 36.0 |
| Seropositive | 9 | 4 (44.4%) | 26 (22 – 44) | 17.0 ± 5.6 | 128.7 ± 33.9 |
| D28** | 6 | 4 (66.7%) | 24.5 (22 – 44) | 17.8 ± 4.7 | 130.5 ± 25.8 |

\*One non-survivor was tested at both days 28 and 42.
\*\* Three seropositive respondents were not tested at 14 days after second dose.

In CoronaVac Phase II trial, there were two times for antibody measurement, 14 or 28 days after second dose (D28 or D42 after first dose).^5^ In our study, there were 2 persons, one non-survivor and one seronegative survivor, measured at both times. Both persons showed increased antibody titer at D42 compared to D28, indicating D42 (28 days after second dose) as more representative time for immune response measurement. However, ANOVA and Bonferroni Post-hoc analysis showed no significant difference of IgG anti-RBD titre between non-survivor at days 28 and 42 as well as seronegative survivor at day 28 (Table 1). Using one sample t-test, the titre of each group was also not significantly different with Phase II trial result.

Figure 1 showed different antibody pattern between seropositive and seronegative survivors. Seropositive survivors were significantly boosted after first dose, while four seronegative survivors remained nonreactive at 14 days after first dose. Antibody titres were higher in seropositive survivor after first dose compared to nonsurvivor and seronegative survivor after second dose. However, anti-RBD titer after first and second dose did not differ significantly in seropositive survivors (Table 2).

**Table 2.** Geometric Mean Titer of post-vaccination IgG anti-RBD between COVID-19 survivors and non-survivors

| Groups | n | GMT [95% CI] |
| --- | --- | --- |
| **Non-survivor\*** |  |  |
| Day 28 | 24 | 32.7 [23.4 – 45.8] |
| Day 42 | 14 | 38.8 [23.0 – 65.2] |
| **Survivor\*\*** |  |  |
| Seronegative D28 | 9 | 41.0 [12.9 – 130.6] |
| Seropositive pre-vaccination | 9 | 114.2 [50.0 – 261.0] |
| Seropositive D14 | 9 | 648.6 [360.7 – 1,166.0] |
| Seropositive D28 | 6 | 845.5 [466.7 – 1,531.8] |
| **CoronaVac Phase II trial<sup>5</sup>** | 117 | 27.6 [22.7 – 33.5] |

\*One non-survivor was tested at both days 28 and 42.
\*\* Three seropositive respondents were not tested at day 28.

**Figure 1.**
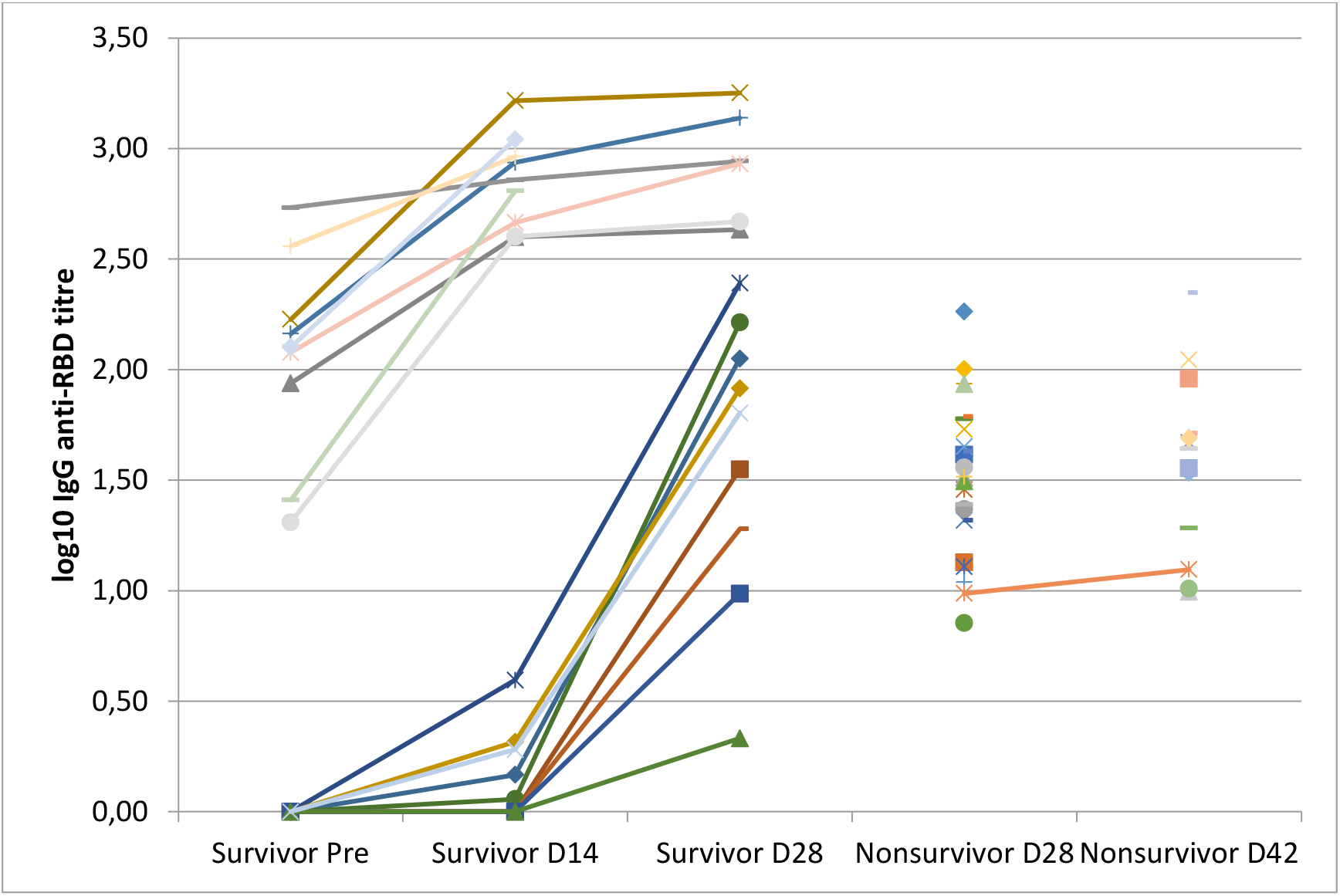
Log10 plot of IgG anti-RBD titer

This preliminary data showed different response pattern between seropositive and seronegative survivors after vaccination with CoronaVac, a whole-inactivated COVID-19 vaccine. We argue that seropositive survivors still need to be vaccinated, at least once. Rise of variants of concern capable of immune escape warrants better immunity response against SARS-CoV-2. Previous study on mRNA vaccine showed vaccinated survivor had increased antibody titre and neutralising activity on both Wuhan Hu-1 and B.1.35.1 strains.^6^ Study on an adenovirus-vectored vaccine, ChAdOx1 nCoV-19, also showed high level of neutralising antibodies towards the SARS-CoV-2 wild type and three variants of concern (B.1.1.7, B.1.35.1, and P1) in survivors receiving a single dose of vaccine.^4^ Whether the same can be observed from whole-inactivated vaccine remains to be observed.

This preliminary study has limitations. We only included small number of respondents, most of them are young healthcare workers. This was due to their prioritisation in the first vaccination phase. However, we plan to include more participant as vaccination eligibility grows. We also only focused on single antibody, IgG anti-RBD, without neutralisation data.

However, CoronaVac study found high positive correlation between IgG anti-RBD titer with neutralising antibody to live SARS-CoV-2 at 14 days (0.80, 95% CI 0.75-0.86) and 28 days (0.85, 95% CI 0.82-0.92) after second dose.^4^

Finally, this study showed that seropositive survivor had significant increase on IgG anti-RBD titer after first CoronaVac vaccine, but second dose did not significantly increase the titer. Therefore, antibody testing (preferably using rapid kit) might be incorporated into the screening process prior to first vaccination dose to enable prioritisation of booster doses for seronegative individuals. This could potentially ease the vaccine supply constrain. A long-term follow-up must be conducted to observe difference in antibody response and persistence.

## Data Availability

-

